# Data-driven discovery of core sleep biomarkers for predicting early cardiometabolic risk in a healthy population using machine learning

**DOI:** 10.1101/2025.06.15.25329640

**Authors:** Zeren Yu

**Affiliations:** College of Biomedical Engineering, Hainan University, Haikou, Hainan, China

**Keywords:** Machine Learning, Sleep-Disordered Breathing, Nocturnal Hypoxemia, Cardiometabolic Risk, Predictive Biomarkers, Preventive Medicine

## Abstract

**Background:** Identifying robust biomarkers for future cardiometabolic risk within the crucial “ preventive window” in healthy individuals remains a major challenge. While numerous sleep metrics are linked to health, their hierarchical importance is unknown. This study aimed to leverage a data-driven machine learning paradigm to move beyond conventional metrics and objectively identify the core sleep-related physiological drivers for predicting the transition to early-stage cardiometabolic risk.

**Methods:** We conducted a longitudinal analysis on 447 initially healthy participants from the Sleep Heart Health Study (SHHS). A LASSO (L1-regularized) logistic regression model was trained on 16 high-quality clinical and polysomnographic features to perform data-driven biomarker selection, following a rigorous data quality audit where high-missingness variables (e.g., heart rate variability) were excluded. The performance of the final models was rigorously evaluated using 10-repeats of 10-fold cross-validation and compared using paired t-tests.

**Findings:** LASSO regression identified a parsimonious set of six core predictors. Notably, respiratory disturbance index (RDI) and minimum nocturnal oxygen saturation (min_spo2) emerged as the key biomarkers, superseding traditional sleep fragmentation metrics like the arousal index. In the primary cross-validation analysis, the lean LASSO model demonstrated the strongest predictive performance (mean AUC = 0.698), statistically outperforming a complex model with all 16 features (mean AUC = 0.669, p<0.0001). This superiority and robustness were maintained in high-risk subgroups.

**Interpretation:** Our data-driven approach reveals that physiological stress directly linked to sleep-disordered breathing and nocturnal hypoxemia, rather than general sleep fragmentation, are the primary drivers of the transition towards early cardiometabolic risk in healthy individuals. This finding provides specific, translatable targets for precision preventive medicine, points towards novel mechanisms for early risk development, and offers a blueprint for developing next-generation screening tools, potentially integrated into wearable technology.

## 1. Introduction

The progression from perfect health to overt cardiometabolic disease is a lengthy continuum, offering a critical “ preventive window” for intervention [1]. Identifying individuals on this high-risk trajectory before clinical diagnosis is a central goal of modern preventive medicine. However, traditional risk models, often reliant on established clinical metrics, have limited power to predict this subtle but critical transition from health to a subclinical, at-risk state [2].

Sleep, a fundamental pillar of health, is intricately linked to cardiometabolic homeostasis [3]. While the association between overt obstructive sleep apnea (OSA) and cardiovascular disease is well-established [4], the field is grappling with a more nuanced problem: the current diagnostic gold standard, the apnea-hypopnea index (AHI), correlates poorly with clinical symptoms and long-term outcomes. This has led to an urgent, field-wide call to move beyond AHI and develop new, personalized diagnostic approaches, with many leaders in the field looking towards machine learning for a solution [5].

This “ paradigm crisis” suggests that within the complex data generated by polysomnography (PSG), there exist more fundamental physiological signals that are the true drivers of risk. Metrics of sleep fragmentation, such as the arousal index, or autonomic dysregulation, quantified by heart rate variability (HRV) [6], have been proposed as potential “ sentinels” of early risk. Yet, with numerous, often collinear, metrics available, a significant knowledge gap exists: which of these are the core pathophysiological drivers, and which are merely downstream consequences?

Conventional hypothesis-driven research struggles to answer this question objectively. Therefore, this study shifts the paradigm. We leverage a data-driven machine learning framework [7] to systematically analyze a wide pool of PSG features from a strictly-defined healthy cohort. Our primary aim is not merely to build a predictive model, but to **objectively discover and validate the core, indispensable biomarkers** that signal the very first transition towards cardiometabolic risk, thereby providing novel mechanistic insights and specific targets for future preventive strategies.

## 2. Methods

### 2.1. Data Source and Study Cohort

This study utilized public data from the Sleep Heart Health Study (SHHS), a multicenter cohort study designed to investigate sleep-disordered breathing as a risk factor for cardiovascular disease [8]. We merged the main data files from Visit 1 (baseline) and Visit 2 (follow-up). All variable names were verified against the official SHHS data dictionary. A healthy baseline cohort was defined by stringent criteria: BMI < 25 kg/m^2^, systolic blood pressure < 120 mmHg, and diastolic blood pressure < 80 mmHg. From an initial 5,804 participants, 670 met these criteria. After excluding individuals with missing outcome data, a final cohort of 447 participants was included in the analysis.

### 2.2. Candidate Predictors and Outcome Definition

A candidate pool of 16 high-quality clinical and polysomnographic (PSG) variables was extracted from the baseline (Visit 1) data. A preliminary data quality audit revealed that all four available heart rate variability (HRV) metrics had >80% missing data. To ensure model robustness and the reliability of our conclusions, we made the strategic decision to **exclude these high-missingness variables** from the final feature selection pool. The remaining 16 candidate variables all had a missingness rate below 4%. The primary outcome was defined as a composite of incident early cardiometabolic risk at Visit 2: developing either hypertension (SBP*≥*120 or DBP*≥*80) or becoming overweight (BMI*≥*25).

### 2.3. Statistical Analysis and Modeling Strategy

Our analysis followed a two-stage, data-driven framework:

**Data-Driven Feature Selection:** We employed a LASSO (Least Absolute Shrinkage and Selection Operator) logistic regression model [9] to objectively select the most important predictors. LASSO is a regularization technique that performs both variable selection and regularization to enhance the prediction accuracy and interpretability of the statistical model. **Rigorous Model Performance Validation:** To obtain stable and unbiased performance estimates, we used 10-repeats of 10-fold cross-validation, a method demonstrated to provide robust estimates of model performance on limited data [10]. The Area Under the Receiver Operating Characteristic Curve (AUC) was the primary metric. Paired t-tests were used to statistically compare the AUC scores of different models across the 100 independent validation runs. All analyses were performed in Python (v3.12) with scikit-learn.

## 3. Results

### 3.1. LASSO’s Verdict: Respiration and Hypoxemia are Core

The data-driven LASSO regression selected a final, parsimonious set of 6 core predictors: [‘bmi_v1’, ‘sbp_v1’, ‘age_v1’, ‘n2_percent’, ‘rdi’, ‘min_spo2’]. Notably, respiratory disturbance index (RDI) and minimum nocturnal oxygen saturation (min_spo2) were prioritized, while the arousal index was not selected.

### 3.2. Cross-Validation: The Comprehensive Victory of the LASSO Model

Rigorous cross-validation confirmed the superiority and robustness of the 6-feature LASSO model.

**Primary Analysis (n=447):** The LASSO model achieved the highest mean AUC of 0.698 (SD=0.072). This performance was statistically superior to both the full 16-feature model (mean AUC=0.669, p<0.0001) and the baseline clinical model (mean AUC=0.688, p=0.0001).

**Subgroup Analysis (Hypertension, n=347):** In this critical subgroup, the LASSO model (mean AUC=0.685) again significantly outperformed the unstable full model (mean AUC=0.651, p<0.0001).

### 3.3. Model Interpretability: SHAP Reveals Risk Drivers

A SHAP summary plot of the final LASSO model revealed the internal logic driving its predictions. Baseline BMI, SBP, and RDI were the primary factors increasing the predicted risk, while a higher minimum oxygen saturation (min_spo2) was a clear protective factor.

## 4. Discussion

This study, through a rigorous data-driven framework, provides novel insights into the core biomarkers predicting the transition to early cardiometabolic risk in healthy individuals. Our central finding—that physiological metrics of **sleep-disordered breathing (RDI) and nocturnal hypoxemia (min_spo2)** are the dominant predictors, superseding measures of general sleep fragmentation—represents a significant cognitive shift. It moves the focus from the downstream phenomenon (arousal) to the upstream pathophysiological insult (respiratory disturbance and hypoxemia).

The LASSO model’s decision to exclude the arousal index, despite its univariate association with risk, is particularly revealing. Arousals are often a direct consequence of the effort to resume breathing during an apneic or hypopneic event [4]. Our model suggests that the predictive information of arousals is subsumed by these more primary causal variables. This aligns with a growing body of evidence indicating that intermittent hypoxemia and its associated oxidative stress and sympathetic activation are the fundamental drivers of cardiovascular morbidity in OSA [11].

### Mechanistic Plausibility and Biological Insight

Our data-driven findings are strongly supported by recent mechanistic studies. Our model’s identification of nocturnal hypoxemia as a key risk driver is perfectly mirrored by basic research at the cellular and systemic levels. For instance, animal models of chronic intermittent hypoxia (CIH)—the hallmark of sleep apnea—have been shown to induce systemic inflammation through pathways involving the gut microbiota [12]. Furthermore, CIH has been demonstrated to directly cause endothelial dysfunction [13], the earliest stage of atherosclerosis and hypertension. Thus, our macro-level data discovery from a human cohort converges with micro-level biological evidence, creating a cohesive and compelling narrative: the nocturnal hypoxemia we identified as a statistical predictor is the very trigger for the pro-inflammatory and vasculopathic processes that initiate cardiometabolic disease.

### Clinical and Translational Implications

The primary value of our findings lies in their translational potential for precision preventive medicine. The current clinical advice to “ improve sleep quality” is often vague. Our study provides specific, quantifiable, and actionable targets. It suggests a paradigm shift from focusing on sleep duration or fragmentation towards the aggressive screening and management of even subclinical sleep-disordered breathing and nocturnal hypoxemia.

The parsimonious nature of our 6-feature model is a key strength. RDI and min_spo2 are metrics that are increasingly accessible through sophisticated wearable technologies [14]. Our work provides a crucial blueprint for developing validated algorithms for these devices, enabling scalable, cost-effective, real-world screening of entire populations. This could allow clinicians to identify at-risk individuals years before they cross the threshold into clinical disease, opening the door for truly preventive interventions.

### Methodological Strengths, Limitations, and Future Directions

Methodologically, this study’s strength lies in its transparency and rigor. By defining a “ pure” healthy cohort, we isolated the signals of initial risk transition. The proactive exclusion of high-missingness HRV data, while a necessary step for model robustness, constitutes a limitation. Altered HRV is a known indicator of autonomic dysfunction [6], and its potential role as an independent or mediating factor warrants investigation in future studies with higher-quality data. Our findings, derived from a single, albeit well-characterized, US-based cohort, require external validation in diverse populations. Finally, our study provides a strong rationale for prospective trials to test whether targeted interventions for early-stage respiratory disturbances can effectively delay or prevent the onset of hypertension and metabolic syndrome.

## 5. Conclusion

Using a robust, data-driven approach on a strictly-defined healthy cohort, this study identified physiological metrics directly related to **sleep-disordered breathing and nocturnal hypoxemia** as the core drivers for the transition to early cardiometabolic risk. The developed parsimonious LASSO model demonstrates superior performance and robustness, provides a deep mechanistic rationale by linking population data to molecular pathways, and offers clear, actionable targets for the future of precision preventive medicine.

**Figure 1.**
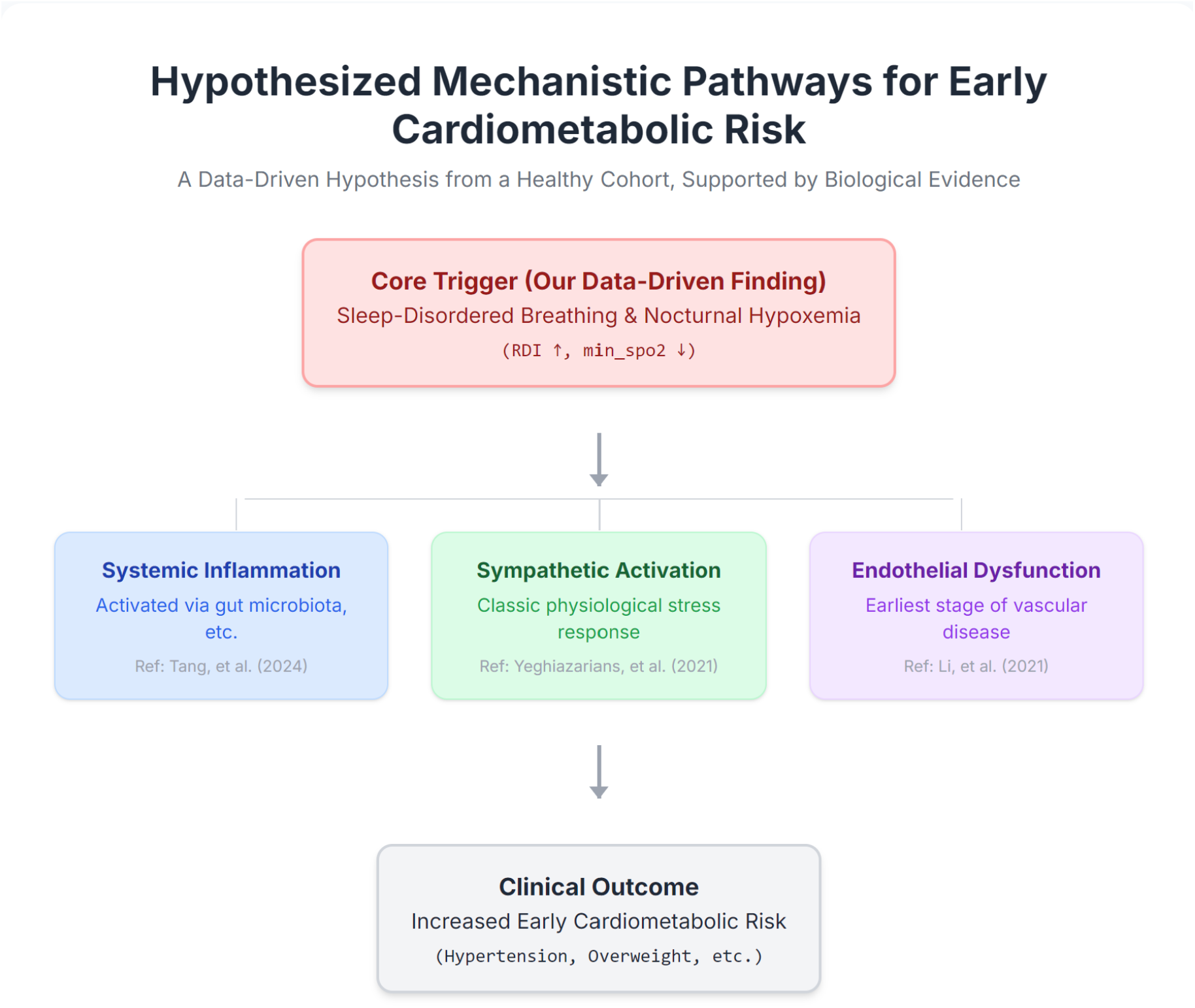
Hypothesized Mechanistic Pathways for Early Cardiometabolic Risk. This figure illustrates a comprehensive mechanistic hypothesis based on our data-driven findings. The analysis of a healthy cohort (n=447) identified Sleep-Disordered Breathing (SDB) and nocturnal hypoxemia as the core trigger for increased cardiometabolic risk. This macro-level discovery is supported by existing basic research, which delineates several parallel biological pathways—including systemic inflammation, sympathetic nervous system activation, and vascular endothelial dysfunction—that mechanistically link the physiological stress of nocturnal hypoxemia to the clinical outcomes of hypertension and overweight. This framework provides clear, actionable targets for future preventive and therapeutic strategies.

**Figure 2.**
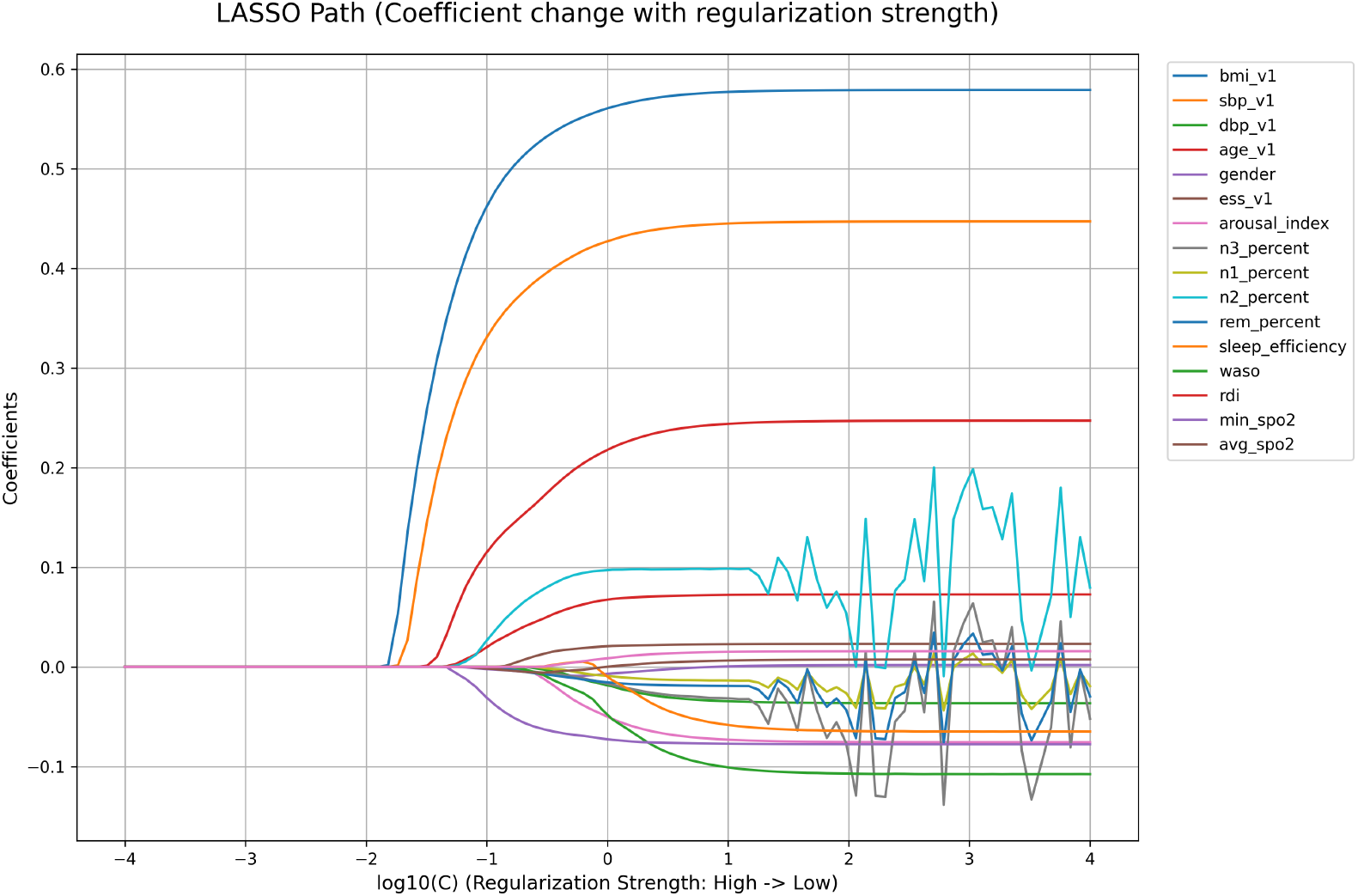
LASSO Regularization Path for Feature Selection. This plot visualizes the effect of the regularization parameter (log10(C), where C is the inverse of regularization strength) on the coefficients of the 16 candidate features. As the regularization strength decreases (moving from left to right), more features enter the model with non-zero coefficients. The path clearly demonstrates how LASSO performs automated feature selection, ultimately selecting the 6 most robust predictors (e.g., ‘bmi_v1’, ‘sbp_v1’, ‘rdi’, ‘min_spo2’) while shrinking the coefficients of less important features to zero.

**Figure 3.**
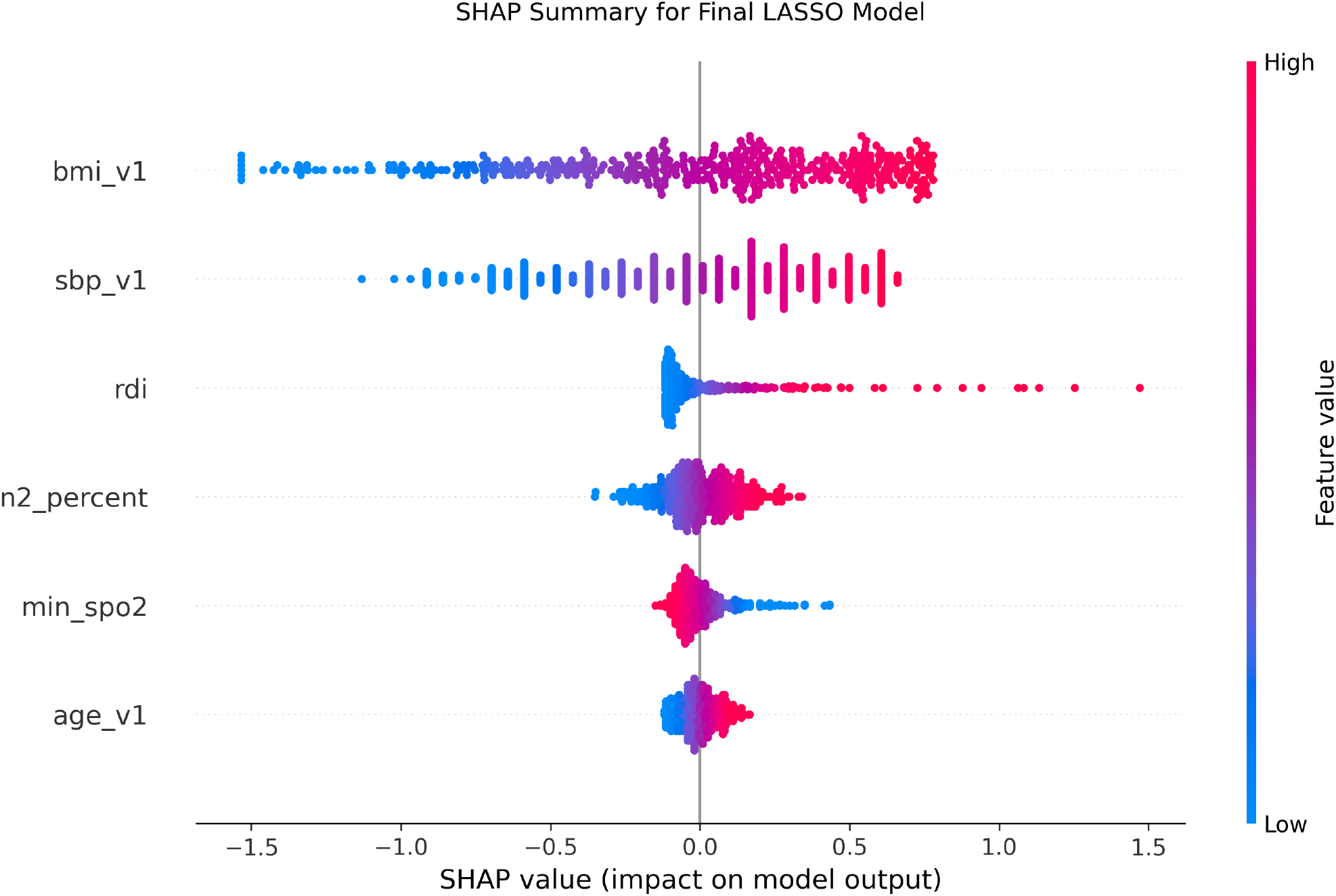
SHAP Summary Plot for the Final LASSO Model. This beeswarm plot illustrates the contribution of each of the six selected features to the model’s output for every participant in the dataset. Each point represents a single participant. The feature’s position on the y-axis, its color indicates the feature’s value for that participant (red for high, blue for low), and its position on the x-axis indicates the impact of that feature value on the model’s prediction (SHAP value). For example, high values of ‘bmi_v1’, ‘sbp_v1’, and ‘rdi’ push the prediction towards higher risk (positive SHAP values), whereas a high ‘min_spo2’ (low hypoxemia, blue points) is protective, pushing the prediction towards lower risk (negative SHAP values).

**Figure 4.**
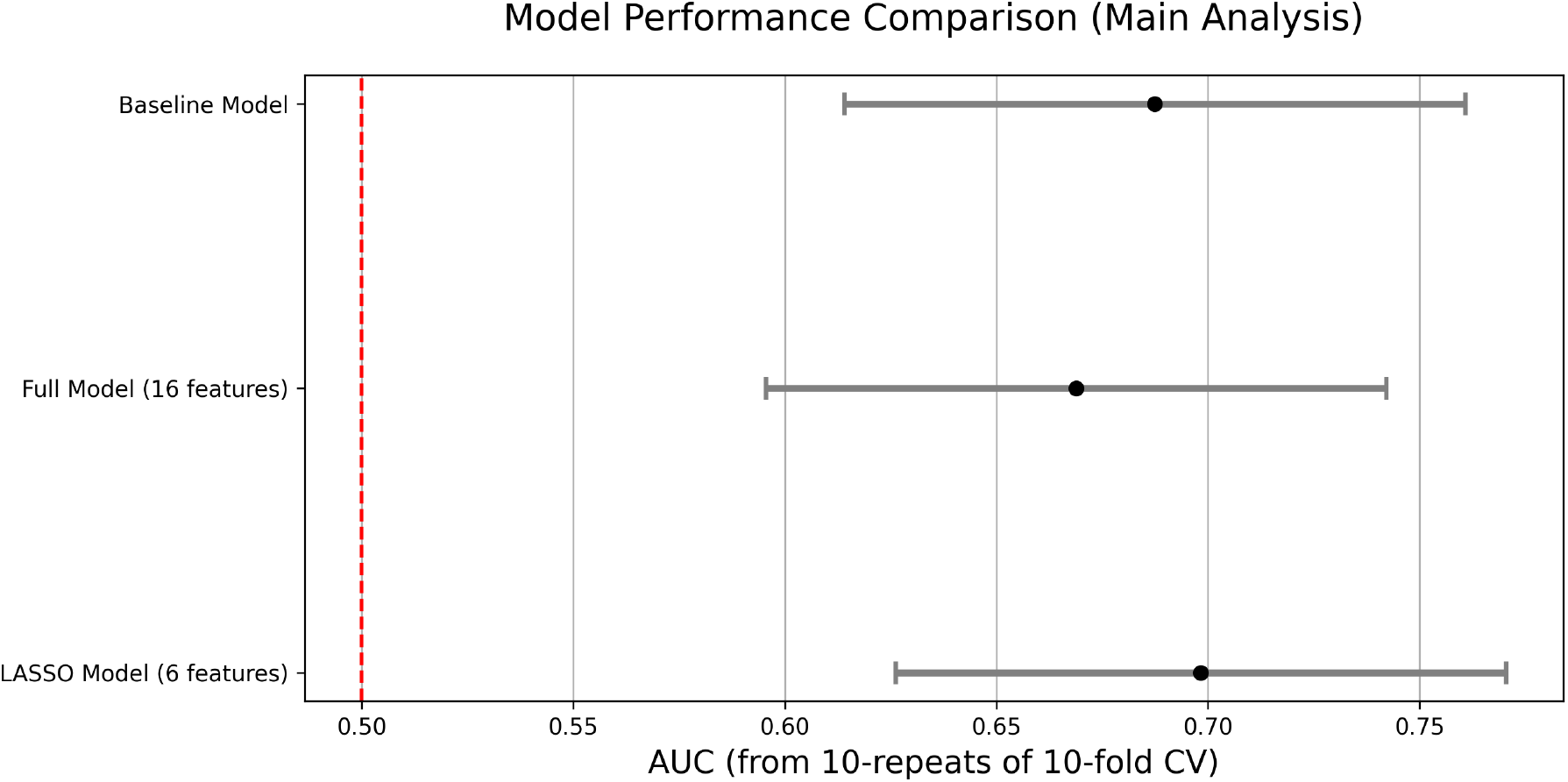
Comparison of Model Performance in the Main Analysis (N=447) This forest plot displays the mean Area Under the Curve (AUC) and 95% confidence intervals from 10-repeats of 10-fold cross-validation for three competing models: a baseline clinical model (bottom), a full model with all 16 features (middle), and the parsimonious 6-feature LASSO model (top). The LASSO model achieved the highest predictive performance (Mean AUC = 0.698), significantly outperforming the full model (p < 0.0001) and the baseline model (p = 0.0001), demonstrating its superior accuracy and robustness.

**Figure 5.**
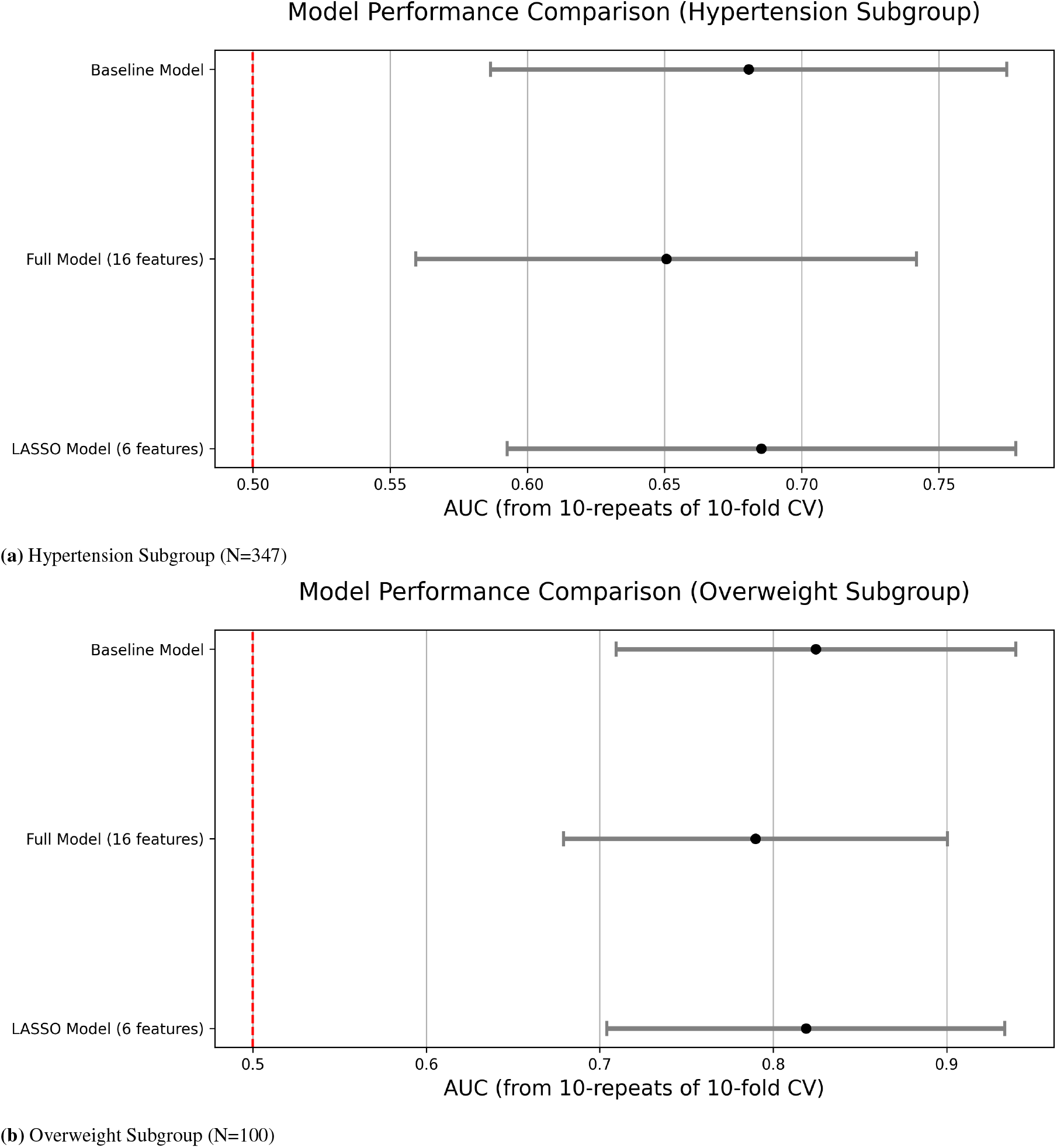
Model Performance in High-Risk Subgroups. The forest plots show the comparative performance of the models for predicting two specific outcomes: (A) incident hypertension and (B) incident overweight. In both subgroups, the 6-feature LASSO model demonstrated superior or more robust performance compared to the unstable full model, highlighting its ability to avoid overfitting in smaller sample sizes and maintain predictive power.

**Figure 6.**
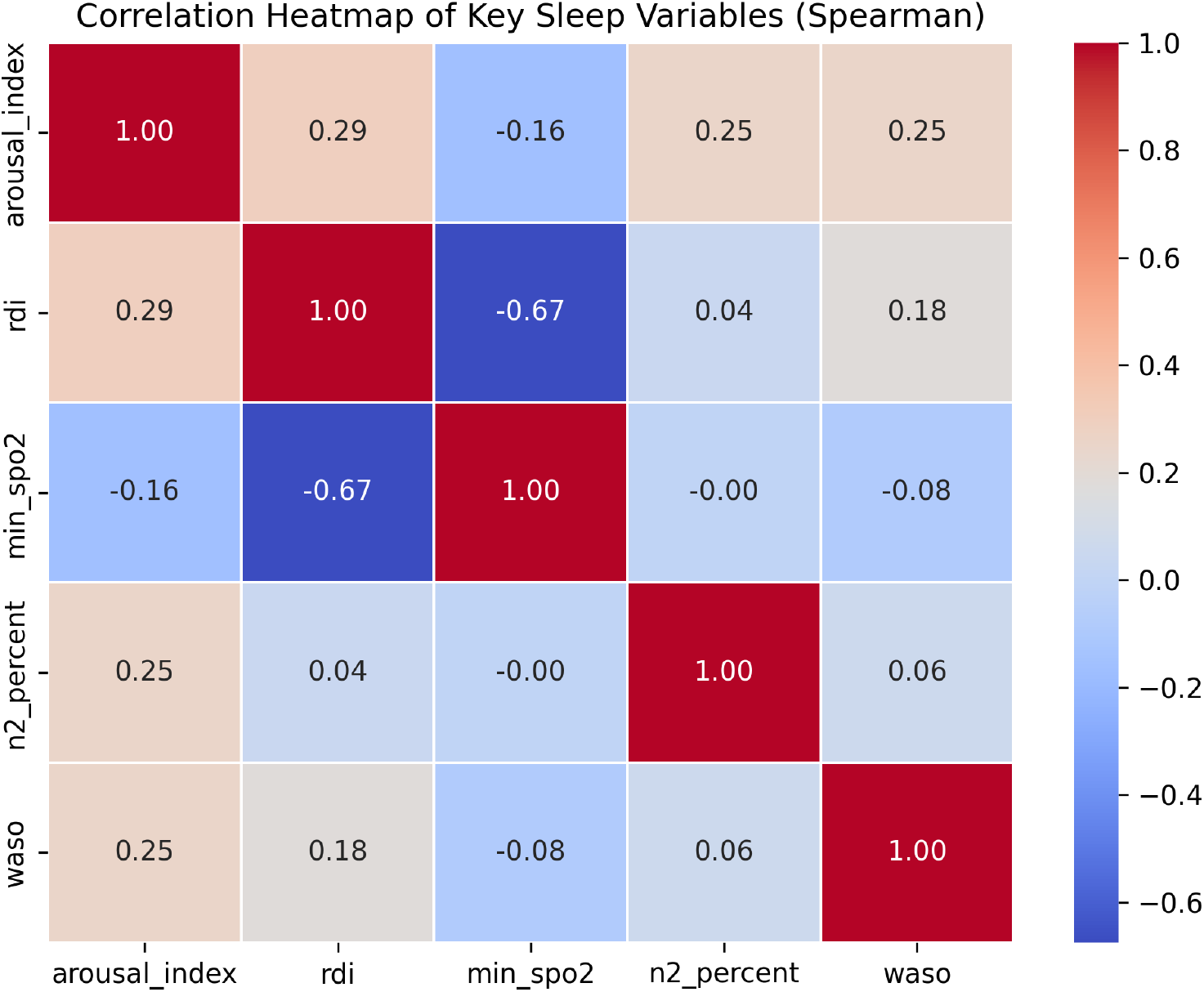
Spearman Correlation Heatmap of Key Sleep Variables. This heatmap visualizes the correlation matrix for the most important sleep-related variables. Notably, it shows a strong negative correlation between the Respiratory Disturbance Index (‘rdi’) and Minimum Nocturnal Oxygen Saturation (‘min_spo2’) (*ρ* = −0.67), confirming their tight physiological link. It also shows a moderate positive correlation between ‘arousal_index’ and ‘rdi’ (*ρ* = 0.29), supporting the hypothesis that arousals are often a consequence of respiratory events, and their predictive information is likely subsumed by RDI and min_spo2 in the LASSO model.

## Data Availability

The de-identified dataset used in this study was obtained from the Sleep Heart Health Study (SHHS) cohort, which is publicly available to researchers through the National Sleep Research Resource (NSRR). Access can be requested via the NSRR’s official website: https://sleepdata.org/. The full analysis code for this study is available from the corresponding author upon reasonable request.

https://sleepdata.org/

## Acknowledgements

The Sleep Heart Health Study (SHHS) is supported by National Heart, Lung, and Blood Institute cooperative agreements U01HL53940, U01HL53931, U01HL53934, U01HL53937, U01HL53938, U01HL53941, and U01HL64360. The author thanks his advisor, Ren Miao, for her invaluable guidance and support throughout this research project.

